# Maternal and fetal HLA heterozygosity in preeclampsia: Insights from a large multi-ancestry pregnancy cohort

**DOI:** 10.64898/2026.06.09.26355260

**Authors:** Chang Cao, Matthew Maher, Jie Hu, Brendan J. Keating, Richard M. Burwick, S. Ananth Karumanchi, G. Larry Maxwell, Camille E. Powe, Thomas F. McElrath, David E. Cantonwine, Norma Serrano, Claudia Colmenares, Juan P. Casas, Richa Saxena, Kathryn J. Gray

## Abstract

Preeclampsia (PE) is a leading cause of maternal and neonatal morbidity, with immune dysregulation at the maternal–fetal interface central to its pathogenesis. The highly polymorphic human leukocyte antigen (HLA) region mediates maternal immune tolerance of the semi-allogeneic fetus, yet the contribution of *HLA* diversity to PE risk remains poorly defined. Whether the *HLA* heterozygote advantage observed in other immune disorders is relevant to PE has not been systematically evaluated. Using data from the multi-ancestry TOPMed Boston–Colombia Collaborative for Adverse Pregnancy Outcomes (n = 12,790; 4,770 PE, 8,020 controls; 10,808 maternal, 1,982 fetal, including 1,848 pairs), we evaluated associations between heterozygosity across eight classical *HLA* loci and PE and four sub-phenotypes, adjusting for genetic ancestry. *HLA* heterozygosity was common across most loci (>80%). No individual maternal *HLA* locus was associated with overall PE; however, heterozygosity across class I loci showed a protective effect in preterm PE (OR=0.82, 95%CI:0.69–0.97), with a similar pattern for *HLA-A* heterozygosity (OR=0.78, 95%CI:0.64–0.96). In contrast, fetal heterozygosity at *HLA-DQB1* was nominally associated with increased risk of PE (OR=1.36, 95%CI:1.03–1.79) and preterm PE (OR=1.73, 95%CI:1.13–2.73). No individual maternal or fetal *HLA* alleles were associated with PE. Maternal–fetal mismatch analysis demonstrated locus-specific associations with preterm PE, including increased risk with *HLA-DQA1* mismatch and reduced risk with *HLA-C* mismatch. These findings highlight distinct maternal and fetal immunogenetic contributions to PE risk and underscore the importance of considering *HLA* diversity—rather than individual alleles alone—in studies of PE etiology.

## Introduction

Preeclampsia (PE) is a severe hypertensive disorder of pregnancy characterized by new-onset hypertension and proteinuria after 20 weeks of gestation. PE affects 2-8% of pregnancies,^1,2^ is a leading cause of maternal and neonatal morbidity and mortality worldwide, and confers a 2-4 fold increase in lifetime risk of cardiovascular disease in affected individuals.^3^ Although clinical symptoms emerge in late gestation, impaired placental remodeling and immune dysregulation in early pregnancy are believed to drive PE pathogenesis.^4,5^ However, the genetic and immunologic factors underlying these abnormalities remain poorly defined, limiting effective strategies for early detection and intervention.

Multiple lines of evidence indicate that maternal–fetal immunogenetic interactions are key regulators of placental development and pregnancy success.^5,6^ Nulliparity and conception using donor sperm or oocytes—contexts that introduce novel paternal antigens—are known PE risk factors.^7,8^ The major histocompatibility complex (MHC), known as the human leukocyte antigen (*HLA*) in humans, spans ∼5 Mb on chromosome 6p21 and encodes >200 immune-related genes.^9,10^ The *HLA* region is divided into three subclasses: class I and class II, which encode antigen-presenting *HLA* molecules, and class III, which contains additional immune genes, including complement components, tumor necrosis factor-α, and MHC class I polypeptide-related sequence A (*MICA*). Class I genes (*HLA-A*, *-B*, *-C*) are expressed on all nucleated cells and present endogenous peptides to CD8+ cytotoxic T cells, whereas class II genes (*HLA-DRB1*, - *DQA1*, -*DQB1*, -*DPA1*, -*DPB1*) are expressed on antigen-presenting cells and present exogenous antigens to CD4+ helper T cells.^11,12^ These classical *HLA* genes exhibit exceptional allelic diversity and high heterozygosity maintained by strong evolutionary selection.^11,12^ Although the *HLA* region harbors more disease associations than any other genomic locus,^13^ its contribution to pregnancy disorders remains poorly characterized.

Pregnancy presents a unique immunologic environment in which cells from two distinct genomes (i.e., maternal and fetal) interact directly at the placental interface. Extravillous trophoblasts (EVT), fetal cells that invade the maternal uterus, express a restricted set of *HLA* genes (*HLA-C, -E, -G*) that interact with killer immunoglobulin-like receptors (*KIR)* on uterine natural killer (NK) cells, thereby contributing to immune tolerance and vascular remodeling.^5,14,15^ The best-characterized interaction–maternal *KIR* AA genotype and fetal *HLA-C2* allele–has been associated with increased risk of PE^16–18^ and recurrent miscarriage^19^ in studies from Europe and sub-Saharan Africa. Mechanistic studies suggest that this pairing reduces NK-cell activation and impairs EVT differentiation and invasion.^20,21^ However, this *HLA–KIR* combination occurs in only a small proportion of PE cases, varies substantially across populations,^16^ and has not been consistently replicated,^22,23^ underscoring the complexity of the immunogenetic landscape of PE.

Beyond specific *HLA–KIR* interactions, the broader contribution of *HLA* genetic variation to PE remains unclear. Genome-wide association studies (GWAS) in predominantly European cohorts have identified two intronic variants in the *HLA* region—rs2442752 in *MICA*^24^ and rs2596471 in *PSORS1C2*^25^—with modest associations with maternal PE risk (odds ratio < 1.1), and neither has been adequately tested in non-European populations. Similarly, studies evaluating maternal–fetal *HLA* compatibility have produced inconsistent results,^23,26–28^ likely due to limited sample sizes (<80 cases) and clinical heterogeneity of PE cases. Together, these observations underscore the need for large, ancestry-diverse cohorts to clarify immunogenetic contributions to PE risk.

Another underexplored aspect of *HLA* in PE is allelic diversity. The extreme polymorphism and high heterozygosity of *HLA* loci are thought to be maintained by pathogen-driven balancing selection, enhancing immune adaptability.^29,30^ The “heterozygote advantage” hypothesis posits that individuals carrying two different *HLA* alleles may present a broader range of antigens, thereby improving immune recognition and host defense.^31–33^ Consistent with this theory, *HLA* heterozygosity has been linked to slower HIV progression,^34,35^ protection against hepatitis B,^36^ and reduced cancer risk.^37–39^ However, its relevance to pregnancy and PE remains largely unexplored. Limited studies have reported lower maternal *HLA-B* heterozygosity in severe PE^40^ and greater fetal *HLA-A* heterozygosity in early-onset PE,^41^ but these findings have not been replicated or extended across all classical *HLA* loci.

The Boston–Colombia Collaborative for Adverse Pregnancy Outcomes (BCC-PREG), a PE-focused study within the National Heart, Lung, and Blood Institute (NHLBI) Trans-Omics for Precision Medicine (TOPMed) program, provides a unique opportunity to address these gaps. This cohort includes ∼13,000 maternal and fetal samples from diverse ancestries, with high-resolution imputation of all classical *HLA* loci. Leveraging these data, we evaluated whether maternal and fetal heterozygosity across *HLA* class I and class II genes is associated with overall PE and its subtypes, and whether allele frequencies and heterozygosity patterns vary by ancestry.

We hypothesized that (a) greater *HLA* heterozygosity would be associated with lower PE risk, reflecting enhanced immune adaptability, and (b) the direction and magnitude of these associations would differ between maternal and fetal genomes and across *HLA* loci, reflecting their distinct roles in immune regulation in pregnancy. We also assessed whether specific *HLA* alleles or maternal–fetal mismatches contribute to PE risk, providing a multidimensional view of *HLA* variation and maternal–fetal immunogenetics in PE.

## Subjects and methods

### Study populations

The BCC-PREG study is composed of five diverse pregnancy cohorts [GenPE (Colombia), LIFECODES (Boston, MA), Burwick/BIDMC (Boston, MA), SPRING (Boston, MA), and Inova (Falls Church, VA)], assembled with the goal of identifying genetic and other molecular determinants underlying PE etiology. Subject characteristics of participants contributing to this analysis (10,808 maternal and 1,982 fetal) are described in **Table 1**.

**Table 1.** Subject characteristics of preeclampsia cases and controls in the BCC-PREG study^a^.

|  | Preeclampsia | Controls |
| --- | --- | --- |
| Maternal samples, N | 3,790 | 7,018 |
| Fetal samples, N | 980 | 1,002 |
| Maternal age (years) | 21.9 (6.4) | 24.7 (8.1) |
| Study cohort, N (%) |  |  |
| GenPE | 3,195 (84.3%) | 3,982 (56.7%) |
| LIFECODES | 306 (8.1%) | 2,530 (36.1%) |
| Burwick/BIDMC | 140 (3.7%) | 220 (3.1%) |
| SPRING | 9 (0.3%) | 150 (2.1%) |
| Inova | 140 (3.7%) | 136 (1.9%) |
| Self-reported ancestry, N (%) |  |  |
| African | 653 (17.2%) | 850 (12.1%) |
| Asian | 31 (0.8%) | 233 (3.3%) |
| European | 310 (8.2%) | 1,784 (25.4%) |
| Hispanic/Latino | 458 (12.1%) | 933 (13.3%) |
| Mixed | 2,178 (57.5%) | 3,004 (42.8%) |
| Others/Unknown | 160 (4.2%) | 214 (3%) |
| Pre-pregnancy BMI (kg/m<sup>2</sup>) | 24.2 (5.7) | 24.2 (6.1) |
| Obesity, N (%) | 324 (8.5%) | 741 (10.6%) |
| Nulliparous, N (%) | 3,441 (90.8%) | 5,405 (77%) |
| Gestational diabetes, N (%) | 47 (1.2%) | 221 (3.1%) |
| Chronic hypertension, N (%) | 102 (2.7%) | 111 (1.6%) |
| PE subtypes, N (%) |  |  |
| Early onset, < 34 weeks | 480 (12.7%) |  |
| Late onset, ≥ 34 weeks | 2,892 (76.3%) |  |
| Preterm, < 37 weeks | 1,232 (32.5%) |  |
| HELLP | 510 (13.5%) |  |
| Gestational age at delivery (weeks) | 36.6 (3.5) | 38.5 (2.6) |
| Preterm Birth, N (%) | 1,232 (32.5%) | 599 (8.5%) |
| Multiple gestation, N (%) | 119 (3.1%) | 186 (2.7%) |
| Birthweight (kg) | 2,627 (783) | 3,115 (533) |
| Fetal sex, N (%) |  |  |
| Male | 1,891 (49.9%) | 3,423 (48.8%) |
| Female | 1,692 (44.6%) | 3,289 (46.9%) |

<sup>a</sup> Values reported as mean (standard deviation) for numerical variables or counts (percentage) for categorical variables

GenPE (International Study of Genetics and Preeclampsia) is a large, multicenter case–control study conducted between 2000 and 2012 across eight cities in Colombia.^42–46^ The study enrolled young, primigravid women without a history of major comorbidities, including diabetes, renal, or cardiovascular disease. A total of 3,702 PE cases and 4,705 normotensive controls were recruited at delivery. Demographic and clinical information—including age, race, socioeconomic status, prenatal care attendance, systolic and diastolic blood pressure, gestational age at delivery, mode of delivery, birth weight, and fetal sex—were collected using standard questionnaires and abstracted from medical records. Participants were predominantly young, of low socioeconomic status, and represented a highly admixed ancestry reflective of the Colombian population. PE diagnosis was standardized across recruitment sites using American College of Obstetricians and Gynecologists (ACOG) criteria.^2^ Maternal and cord blood samples were collected at delivery and used for DNA extraction and subsequent whole genome sequencing (WGS).

LIFECODES is a pregnancy biorepository established in 2006 in the Maternal–Fetal Medicine Division at Brigham and Women’s Hospital (BWH) in Boston, MA.^47–52^ Eligibility criteria included initiation of prenatal care before 16 weeks of gestation, age ≥ 18 years, and planned delivery at BWH. Maternal urine and plasma samples were collected at enrollment (∼ 11 weeks of gestation) and at follow-up visits (∼ 26 and 35 weeks of gestation). Sociodemographic characteristics and medical history were collected via standardized questionnaires, and pregnancy outcomes were abstracted at delivery. Adverse pregnancy outcomes, including PE and preterm birth were reviewed and validated by two physicians. Maternal DNA was extracted from all available buffy coat samples, and fetal DNA was obtained from cord blood or placental samples when available, for inclusion in the BCC-PREG WGS dataset.

The Burwick/BIDMC cohort consists of PE case–control studies conducted at BWH (2011–2013) and Beth Israel Deaconess Medical Center (2008–2014) in Boston, MA, recruiting individuals with PE at the time of presentation and normotensive controls.^53–55^ Maternal and cord blood/placenta samples underwent DNA extraction for WGS in BCC-PREG.

The Study of Pregnancy Regulation of Insulin and Glucose (SPRING) is a longitudinal study of glucose metabolism in 166 pregnant individuals at high risk for gestational diabetes recruited from Massachusetts General Hospital and the general population in Boston, MA between 2016 and 2021.^56^ Participants were enrolled in the first trimester and completed two oral glucose tolerance tests during pregnancy and one postpartum. DNA was extracted from maternal buffy coats for WGS.

Inova participants were from the “Genomic Correlations to Childhood Health Outcomes” study conducted within the Inova Health System (Falls Church, VA) between 2011 and 2013.^57^ The parent study enrolled 791 family trios to investigate the genetic and molecular determinants of preterm birth and related disorders. A subset of mothers with PE and matched normotensive controls, along with their offspring, were included in BCC-PREG, with DNA extracted from maternal and cord blood for WGS.

The BCC-PREG study was approved by the Institutional Review Board (IRB) at the Mass General Brigham. All contributing studies were approved by their respective IRBs, and written informed consent was obtained from all participants.

### Phenotyping and PE definitions

PE cases were defined as new-onset hypertension (SBP ≥ 140 mmHg or DBP ≥ 90 mmHg on two occasions, at least 4 hours apart) and proteinuria (24-hour urine protein ≥ 300 mg or spot protein/creatinine ratio > 0.3) after 20 weeks of gestation according to ACOG guidelines.^2^ PE subtypes were defined by the following criteria: early-onset (EOPE) with delivery < 34 weeks of gestation; late-onset (LOPE) with delivery ≥ 34 weeks of gestation; preterm PE with delivery < 37 weeks of gestation; and Hemolysis Elevated Liver enzyme Low Platelets (HELLP) syndrome, defined by liver enzymes (aspartate aminotransferase (AST) or alanine aminotransferase (ALT)) > 2 × the upper limit of normal and platelet count < 100 × 10^9^/L. For this genetic analysis, an individual was considered a case if they had a diagnosis of PE in either current or any previous pregnancy. Controls were normotensive pregnant women with no prior history of PE.

### Whole genome sequencing (WGS)

WGS was generated through the NHLBI TOPMed program for 14,262 DNA samples in the BCC-PREG study (12,179 maternal and 2,084 fetal samples). Sequencing was performed at the Broad Institute (Cambridge, MA), one of the seven TOPMed genomic centers, using 150 bp paired-end reads at a mean depth of 38x.^58^ Detailed descriptions of TOPMed WGS protocols, alignment, variant calling, harmonization, and quality control procedures have been published previously.^58,59^ All BCC-PREG genotype data used in this study were obtained from TOPMed Freeze 12, which consists of jointly called variants from WGS data by the TOPMed Informatics Research Center (IRC) using the GRCh38 reference genome. TOPMed Freeze 12 contains joint calls from 217,936 samples on 961.9 million SNPs and 79.1 million Indels.

We performed additional sample-level quality control on the genotype data, including evaluation of duplicate samples, pedigree inconsistencies, and sex discrepancies. Following quality control, a total of 12,790 samples (10,808 maternal and 1,982 fetal) were retained for analysis. Principal components (PCs) of genetic ancestry were computed for all samples using PLINK2.

### *HLA* imputation

High-resolution (four-digit) alleles for eight classical *HLA* genes (class I: *HLA-A*, *-B*, *-C*; class II: *HLA-DRB1*, *-DQB1*, *-DQA1*, *-DPB1*, *-DPA1*) were imputed from genotype data using a multi-ancestry *HLA* imputation reference panel^60^ comprising 20,349 individuals from Admixed African, East Asian, European, and Latino ancestry. Imputation was performed through the Michigan imputation server (MIS) using Minimac4.^61^ The *HLA* imputation procedure and its benchmarked performance across diverse populations, including those in the 1000 Genomes Project, have been described in detail by Sakaue et al.^9^ Since becoming publicly available on the MIS in 2022, this multi-ethnic *HLA* reference panel has been applied in multiple independent studies to investigate *HLA*-disease associations, including analyses in Admix Africans,^62^ Europeans,^63,64^ and multi-ancestry cohorts encompassing European, Admix American, African, and Asian populations.^65–67^ As an additional quality control step, Mendelian consistency of imputed *HLA* genotypes was assessed in paired maternal–fetal samples (n = 1,848 pairs). No maternal–fetal pairs exhibited more than one Mendelian error across the eight *HLA* loci, and 95% of pairs showed complete Mendelian consistency, supporting the overall quality of the imputed *HLA* data.

### *HLA* heterozygosity

For each sample, heterozygosity was calculated for: 1) each *HLA* locus, 2) class I loci, and 3) class II loci. Consistent with previous studies,^34,37,38,68^ we considered a sample heterozygous at a specific locus if the two four-digit alleles were distinct. Full heterozygosity at class I loci was defined as having all six alleles across the three loci be distinct. Full heterozygosity at class II loci was defined as having all ten alleles across the five loci be distinct.

## Statistical Analyses

### *HLA* locus heterozygosity and PE risk

To evaluate the association between *HLA* heterozygosity and risk of PE, we performed multivariate logistic regression for each *HLA* locus as well as for composite measures of class I and class II heterozygosity. All models were adjusted for maternal age, study cohort, and ten genetic ancestry PCs to account for population structure. Associations were examined for overall PE and for PE subtypes, including EOPE, LOPE, preterm PE, and HELLP syndrome. Analyses were conducted separately in maternal samples (n = 10,808) and fetal samples (n = 1,982). Fetal models were additionally adjusted for fetal sex. Maternal models were not adjusted for fetal sex because fetal sex of the pregnancy was missing for 513 maternal cases; exclusion of this variable allowed maximization of sample size and power in maternal analyses. To account for multiple testing across the eight locus-specific and two composite class I and class II heterozygosity measures, *p* values were adjusted using the Benjamini–Hochberg false discovery rate (FDR) procedure, with FDR-adjusted *p* < 0.05 considered statistically significant. For fetal HELLP analyses (n = 15 cases), standard logistic regression exhibited complete separation for *HLA-A* and *HLA-DPB1* heterozygosity. Therefore, Firth penalized logistic regression^69^ was applied across all *HLA* loci to obtain bias-reduced odds ratios and confidence intervals.

### Individual *HLA* alleles and PE risk

To investigate the association of individual *HLA* alleles with PE risk, we restricted analyses to alleles with a frequency greater than 1%, a commonly used cut-off for *HLA* allele–disease association studies.^38,70^ Of the 452 imputed alleles in BCC-PREG, 130 alleles met this criterion and were included in the analysis. For each sample, allele dosage was coded as 0, 1, or 2 as previously described^38^ and tested in multivariate logistic regression analysis with PE or PE subtypes as outcomes. Models were adjusting for maternal age, study cohort, and ten genetic ancestry PCs; fetal sex was additionally included as a covariate in fetal analyses. To ensure stable estimation, alleles with fewer than five carriers among cases or few than five carriers among controls for a given outcome were not tested. FDR-adjusted *p* values were calculated using the Benjamini-Hochberg procedure to account for multiple testing within each outcome. Significance was defined as FDR-adjusted *p* value < 0.05.

### Maternal and fetal *HLA* mismatch

We counted the number of mismatched alleles in 1,848 maternal and fetal pairs (944 PE and 904 control) at each *HLA* locus using similar definitions from previous maternal–fetal *HLA* mismatch analyses.^26,28,71^ A mismatch count of 0 at a given locus indicates that the mother and fetus share both alleles. If one allele differs between the mother and fetus, the mismatch count is 1. Mismatch counts were not calculated if either the mother or fetus was missing one or more alleles for a given locus. We also summed the total mismatched allele counts across all 8 *HLA* loci for each pair. The difference in total mismatched allele counts between PE and control pairs was evaluated by a two-sample t-test. The association between *HLA* mismatch and PE risk was tested for each *HLA* locus, controlling for maternal age, study, fetal sex, and ten genetic ancestry PCs. FDR-adjusted *p* values were calculated to account for multiple testing across *HLA* loci. FDR-adjusted *p* values < 0.05 were considered significant for the mismatch and PE risk associations.

### Subgroup analysis

To assess the robustness of the observed associations and to account for differences in sample size across the five BCC-PREG studies, subgroup analyses were performed by geographic region, grouping samples into US-based cohorts (LIFECODES, Inova, Burwick/BIDMC, and SPRING) and the Colombia-based cohort (GenPE). In addition, to evaluate potential ancestry-specific effects, analyses were also stratified by the three major genetic ancestry groups (Admixed American (AMR), European (EUR), and African (AFR)) as inferred from genetic data.

All statistical analyses were performed using RStudio (version 2023.12.1.402).^72^

## Results

### Subject Characteristics

Subject characteristics of the BCC-PREG study participants are summarized in **Table 1**. Participants had a mean age of 23.8 years and a mean BMI of 24.2 kg/m². The majority of samples in BCC-PREG originated from the Colombia-based GenPE study (n = 7,177: 66.4% of all samples; 87.1% of cases and 55.8% of controls), followed by the Boston-based LIFECODES study (n = 2,836, 26.2%). The remaining 7.4% of samples (n = 795) came from three other US-based cohorts (Inova, Burwick/BIDMC, SPRING). Based on self-reported race/ethnicity, most participants identified as mixed race (47.9%), followed by White (19.4%) and Black (13.9%). Genetic ancestry inference broadly corroborated self-reported classifications, with the majority of samples exhibiting Admixed American (AMR) ancestry (63.5%), followed by European (EUR; 18.7%) and African (AFR; 12.8%) ancestries. The distribution of self-reported race/ethnicity across inferred genetic ancestry groups (AMR, AFR, EUR, EAS, MID, and SAS) is provided in **Table S1**. Consistent with known PE risk factors, PE cases, compared to controls, were more frequently nulliparous (91% vs. 77%), delivered earlier (36.6 vs. 38.5 weeks), had a higher proportion of preterm births (32.5% vs. 8.5%), and lower birthweights (2,627 vs. 3,115 g). Fetal sex and multiple vs. singleton pregnancies did not differ between cases and controls.

### *HLA* Heterozygosity in BCC-PREG

As expected, all eight classical *HLA* loci showed high levels of heterozygosity (> 80% for all loci except *HLA-DPA1*) (**Figure 1**). These values are consistent with reports from UK Biobank, FinnGen,^38^ Ashkenazi Jewish and Arab populations,^37^ and other major global ancestries (AMR, EUR, AFR),^73^ reflecting the well-established evolutionary maintenance of *HLA* diversity. The lower heterozygosity at *HLA-DPA1* (49%) also aligns with patterns reported in other populations. A large proportion of participants were fully heterozygous across all class I loci (83%) and class II loci (36%). Heterozygosity rates were comparable across the four US studies; however, when participants were stratified by geographic region, the Colombian cohort exhibited significantly higher heterozygosity at six of the eight *HLA* loci, with the exceptions of *HLA-DQA1* and *HLA-DQB1* (**Table S2**). Similarly, heterozygosity rates differed significantly across major genetic ancestry groups (EUR, AMR, AFR), with individuals of European ancestry exhibiting lower heterozygosity at most loci except for *HLA-DQA1* and *HLA-DQB1* (**Table S3**). Together, these findings highlight substantial geographic- and ancestry-related variation in *HLA* heterozygosity and underscore the importance of accounting for population structure in pooled analyses and conducting subgroup analyses. In fetal samples (n = 1,982), heterozygosity patterns closely mirrored those of maternal samples, with all loci demonstrating high heterozygosity (> 80%) except *HLA-DPA1* (52%) (**Figure 1**). Similarly, most fetuses were fully heterozygous across class I loci (82%), and 39% were heterozygous across all class II loci.

**Figure 1.**
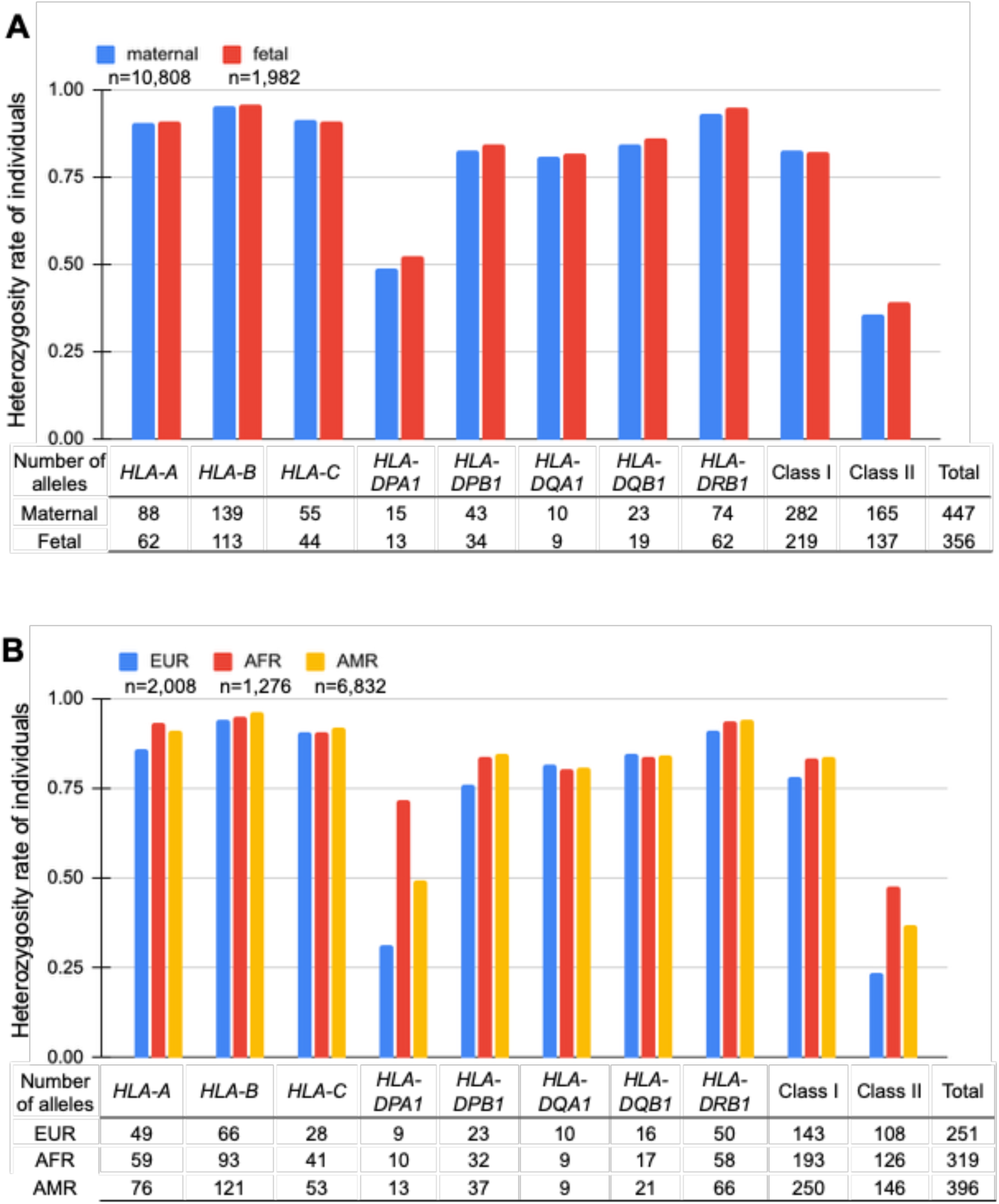
*HLA* heterozygosity among BCC-PREG study participants by maternal and fetal status (**A**) and by major genetic ancestry (**B**).

### Maternal *HLA* Heterozygosity and PE risk

We next examined whether maternal heterozygosity at each *HLA* locus was associated with risk of PE and PE subtypes (EOPE, LOPE, preterm PE, HELLP) using logistic regression adjusted for age, study, and genetic ancestry. In addition to locus-specific analyses, we assessed whether full heterozygosity across *HLA* class I or class II loci was associated with PE risk. None of the individual *HLA* loci were independently associated with overall PE risk (**Figure 2A**). Notably, point estimates for all *HLA* class I loci were directionally consistent with reduced PE risk. This trend was most pronounced for full heterozygosity across class I loci (OR = 0.90, 95% CI 0.81–1.01), although the association did not reach significance (*p* = 0.08; FDR-adjusted *p* = 0.77).

**Figure 2.**
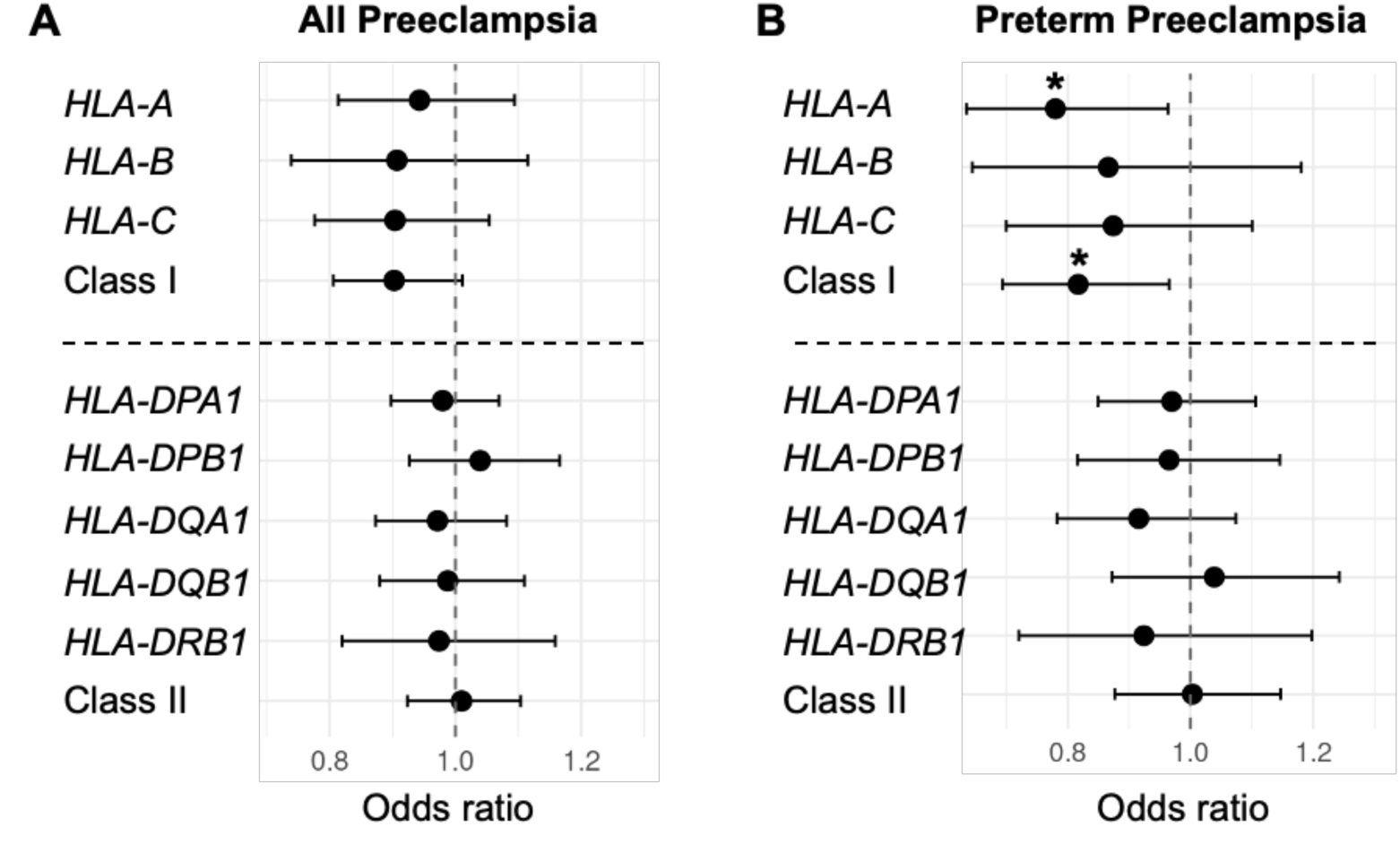
Associations of maternal heterozygosity across 8 classical *HLA* loci and *HLA* class I and class II with risk of preeclampsia (A) and preterm preeclampsia (B). All models adjusted for maternal age, study, and genetic ancestry \**p* value < 0.05 before FDR-correction

In subgroup analyses of preterm PE (n = 1,232), heterozygosity at *HLA-A* (OR = 0.78, 95%CI: 0.64–0.96; *p* = 0.02; FDR-adjusted *p* = 0.11) and full heterozygosity across all class I loci (OR = 0.82, 95%CI:0.69–0.97; *p* = 0.02; FDR-adjusted *p* = 0.11) showed nominal evidence of reduced risk, although neither association remained statistically significant after FDR correction (**Figure 2B**). Despite this, both associations demonstrated a consistent protective direction of effect, supporting a potential role for maternal *HLA* class I heterozygosity in preterm PE risk. To assess potential geographic or ancestry-specific effects, we examined these associations separately in US (n = 268) and Colombian (n = 964) preterm PE cases. Directionally consistent protective effects for *HLA-A* heterozygosity and full class I heterozygosity were observed in both groups; however, statistical significance was reached only in the Colombian cohort, likely reflecting limited power in the smaller US subgroup (**Table S4**). Similarly, analyses stratified by genetic ancestry (EUR, AMR, AFR) revealed consistent protective effects (OR < 1) of *HLA-A* and full class I heterozygosity across all three ancestry groups, although none reached statistical significance (**Table S5**). No associations were observed between maternal *HLA* heterozygosity and other PE subtypes, including EOPE, LOPE, or HELLP (**Table S6**).

### Fetal *HLA* Heterozygosity and PE risk

We next evaluated whether fetal *HLA* heterozygosity was associated with PE risk among 1,982 fetal samples (980 cases and 1,002 controls). Heterozygosity at the fetal *HLA-DQB1* locus showed a nominal association with increased PE risk (OR = 1.36, 95%CI:1.03–1.79, *p* = 0.03, FDR-adjusted *p* = 0.28) (**Figure 3A**). A similar but stronger association was observed in preterm PE cases (n = 279) (OR = 1.73, 95%CI:1.13–2.73, *p* = 0.01, FDR-adjusted *p =* 0.12) (**Figure 3B**).

**Figure 3.**
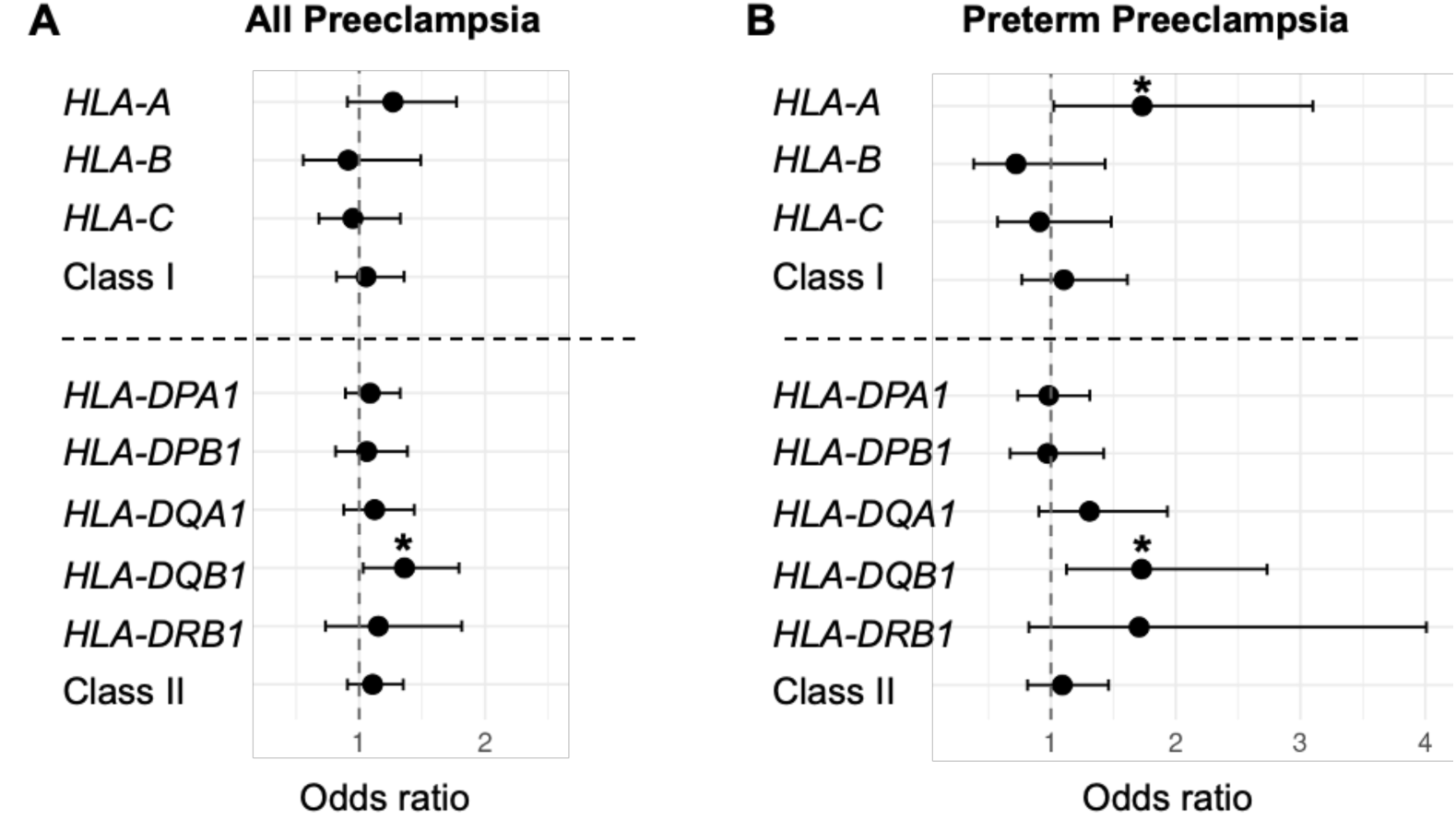
Associations of fetal heterozygosity across 8 classical *HLA* loci and *HLA* class I and class II with risk of preeclampsia (A) and preterm preeclampsia (B). All models adjusted for maternal age, study, fetal sex, and genetic ancestry \**p* value < 0.05 before FDR-correction

Heterozygosity at fetal *HLA-A* also showed a nominally positive association with preterm PE (OR = 1.73, 95%CI:1.02–3.10, *p* = 0.04, FDR-adjusted *p =* 0.21). No other fetal *HLA* loci showed association with PE or PE subtypes (**Table S7**).

### *HLA* Alleles in BCC-PREG

Large *HLA* reference datasets, including the Catalog of Common, Intermediate, and Well-Documented (CIWD) *HLA* alleles, as well as whole-genome resources such as gnomAD, have demonstrated substantial geographic- and ancestry-related variation in *HLA* allele frequencies, particularly among low-frequency and rare alleles.^29,74–77^ Consistent with these patterns, 103 of the 130 *HLA* alleles examined in BCC-PREG showed significant frequency differences (FDR-adjusted p < 0.05) between US-based participants (n = 3,631; predominantly EUR ancestry, 55%) and Colombian participants (n = 7,177; predominantly AMR ancestry, 87%). Despite these absolute differences, allele frequencies were highly correlated between the two groups (Pearson’s r = 0.91, *p* = 5.4 × 10^-52^; **Figure 4**), likely reflecting our focus on alleles with frequencies greater than 1%, which are broadly distributed across populations. Larger studies will be needed to further characterize population-specific patterns for lower-frequency *HLA* alleles in BCC-PREG.

**Figure 4.**
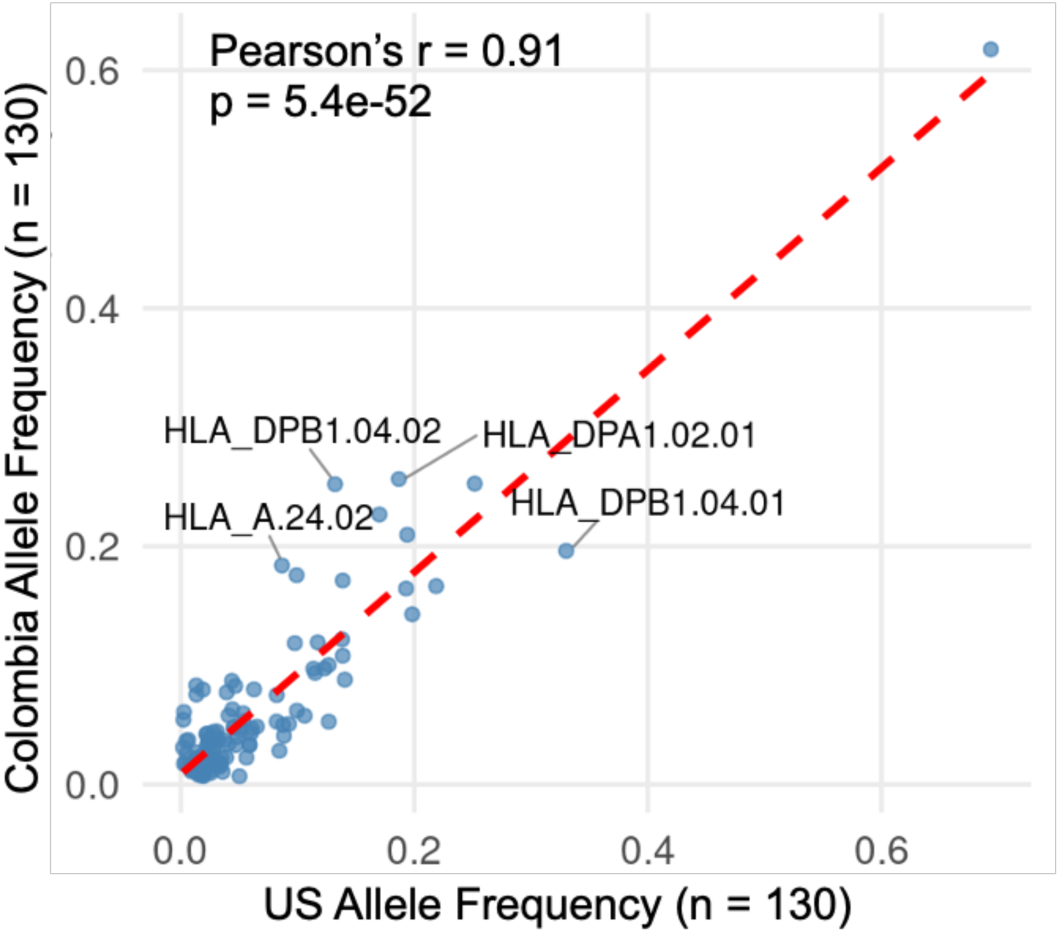
Comparison of *HLA* allele frequencies between the US and Colombian participants in the BCC-PREG study. Scatter plot showing the correlation of allele frequencies for 130 *HLA* alleles between US (n = 3,631) and Colombian (n = 7,177) participants. Allele frequencies were highly correlated between the two geographic groups (Pearson’s r = 0.91, *p* = 5.4e-52). Labeled alleles represent those with standardized residuals > 3 from the fitted correlation line.

As a final quality-control assessment of *HLA* imputation performance, we compared allele frequencies derived from imputed *HLA* genotypes in BCC-PREG with those reported in the CIWD *HLA* allele database,^77^ which aggregates allele frequency data for multiple classical *HLA* genes from approximately eight million individuals across diverse donor registries. For this comparison, we extracted ancestry-specific allele frequencies for the three major genetic ancestry groups represented in BCC-PREG (EUR, AFR, and AMR) for classical *HLA* genes available in CIWD (*HLA-A*, *HLA-B*, *HLA-C*, *HLA-DPB1*, *HLA-DQB1*, and *HLA-DRB1*). Because CIWD does not define AMR as a distinct category, allele frequencies of the Hispanic group representing South or Central America/Hispanic/Latino populations were used as an approximate reference for BCC-PREG AMR samples. Allele frequencies in BCC-PREG were highly correlated with those reported in the corresponding CIWD reference populations across all three ancestry groups (Pearson’s r > 0.93 within each ancestry; **Figure S1**), supporting the accuracy of *HLA* imputation using the multi-ancestry reference panel.

### Maternal and Fetal *HLA* Alleles and PE risk

No individual *HLA* alleles were significantly associated with risk of PE or its subtypes in either maternal or fetal analyses after correction for multiple testing. The ten alleles with the smallest p-values from maternal and fetal analyses are summarized in **Table 2**, and complete association results for all tested alleles are provided in **Tables S8** and **S9**.

**Table 2.** Top ten *HLA* alleles nominally associated with preeclampsia (PE) or four PE subtypes in maternal and fetal analyses^a^.

|  | Allele | Outcome | Odds ratio | Lower 95%CI | Upper 95%CI | *p* | adj_*p*<sup>b</sup> |
| --- | --- | --- | --- | --- | --- | --- | --- |
| maternal | *HLA_C\*16:01* | Early onset PE | 1.547 | 1.192 | 2.008 | 0.001 | 0.132 |
| maternal | *HLA_DRB1\*04:02* | PE | 1.608 | 1.202 | 2.152 | 0.001 | 0.098 |
| maternal | *HLA_DPB1\*02:01* | PE | 1.163 | 1.060 | 1.277 | 0.002 | 0.098 |
| maternal | *HLA_C\*04:01* | Late onset PE | 1.153 | 1.055 | 1.259 | 0.002 | 0.178 |
| maternal | *HLA_DRB1\*04:02* | Late onset PE | 1.609 | 1.169 | 2.214 | 0.003 | 0.178 |
| maternal | *HLA_DQB1\*04:02* | Late onset PE | 0.833 | 0.735 | 0.944 | 0.004 | 0.178 |
| maternal | *HLA_DRB1\*04:11* | Preterm PE | 0.585 | 0.405 | 0.847 | 0.004 | 0.325 |
| maternal | *HLA_DRB1\*04:11* | PE | 0.735 | 0.593 | 0.910 | 0.005 | 0.181 |
| maternal | *HLA_C\*04:01* | PE | 1.121 | 1.034 | 1.216 | 0.006 | 0.181 |
| maternal | *HLA_B\*44:02* | Early onset PE | 1.607 | 1.143 | 2.259 | 0.006 | 0.409 |
| fetal | *HLA_DQB1\*06:09* | PE | 2.621 | 1.434 | 4.787 | 0.002 | 0.225 |
| fetal | *HLA_DPA1\*02:02* | Early onset PE | 2.548 | 1.403 | 4.626 | 0.002 | 0.148 |
| fetal | *HLA_DQB1\*06:09* | Early onset PE | 2.612 | 1.394 | 4.893 | 0.003 | 0.353 |
| fetal | *HLA_B\*15:01* | Early onset PE | 3.150 | 1.383 | 7.174 | 0.006 | 0.220 |
| fetal | *HLA_C\*02:02* | Early onset PE | 2.394 | 1.222 | 4.691 | 0.011 | 0.256 |
| fetal | *HLA_DQB1\*06:09* | Preterm PE | 2.786 | 1.228 | 6.320 | 0.014 | 0.725 |
| fetal | *HLA_A\*29:02* | Preterm PE | 1.727 | 1.088 | 2.741 | 0.021 | 0.725 |
| fetal | *HLA_DQB1\*06:03* | Preterm PE | 1.687 | 1.076 | 2.643 | 0.023 | 0.725 |
| fetal | *HLA_B\*35:03* | Preterm PE | 2.349 | 1.124 | 4.912 | 0.023 | 0.725 |
| fetal | *HLA_DRB1\*13:02* | PE | 1.409 | 1.035 | 1.918 | 0.029 | 0.922 |

<sup>a</sup> All models are controlled for maternal age, study, and genetic ancestry
<sup>b</sup> *p* values are FDR-adjusted for multiple testing across *HLA* alleles using the Benjamini–Hochberg method

### Maternal-fetal *HLA* Mismatch and PE Risk

Finally, we evaluated whether maternal-fetal mismatch in any *HLA* locus affects PE risk in the 1,848 maternal-fetal pairs (944 PE and 904 control) (**Table S10**). None of the *HLA* loci were significantly associated with PE risk after adjusting for maternal age, fetal sex, study, and genetic ancestry; mismatch in *HLA-DQA1* showed a nominal association with PE risk (OR = 1.40, 95%CI:1.09–1.80, *p* = 0.008, FDR-adjusted *p* = 0.06). In subgroup analysis of preterm PE, mismatch in *HLA-DQA1* was significantly associated with increased risk (OR = 1.74, 95%CI:1.16–2.60, *p* = 0.007, FDR-adjusted *p* = 0.03) while mismatch in *HLA-C* was associated with lower risk (OR = 0.54, 95%CI:0.35–0.85, *p* = 0.007, FDR-adjusted *p* = 0.03). Associations of PE with maternal–fetal mismatch across all *HLA* loci are provided in **Table S11**. In addition to individual *HLA* locus, we also counted the total mismatched loci for each maternal–fetal pair and found no difference in the total mismatch count between cases and controls (6.9 ± 1.3 vs. 6.9 ± 1.4, *p* = 0.94; **Table S10**).

## Discussion

In this large, ancestrally diverse pregnancy cohort, we provide a comprehensive assessment of maternal and fetal *HLA* heterozygosity across all eight classical *HLA* loci in relation to PE. We observed that maternal heterozygosity across *HLA* class I loci was associated with reduced risk of PE—particularly preterm PE—whereas fetal heterozygosity at the class II locus *HLA-DQB1* was associated with increased PE risk, with effects most pronounced in preterm PE. These associations were robust to adjustment for genetic ancestry, although some did not remain statistically significant after correction for multiple testing across loci. In contrast, no individual *HLA* alleles were significantly associated with PE or its subtypes, and maternal–fetal *HLA* mismatch showed only modest, locus-specific effects limited to preterm PE. Collectively, these findings suggest that overall *HLA* diversity, rather than specific alleles or simple compatibility patterns, may influence PE susceptibility through distinct maternal and fetal mechanisms.

The protective association of maternal *HLA* class I heterozygosity is consistent with the heterozygote advantage hypothesis, in which carrying divergent *HLA* alleles broadens antigen presentation and enhances immune adaptability. In pregnancy, greater maternal *HLA* diversity may promote a more balanced immune state that preserves tolerance toward the semi-allogeneic fetus while maintaining effective immune surveillance. Our findings extend earlier observations linking lower maternal *HLA-B* heterozygosity to severe PE^40^ and suggest that maternal class I diversity may protect against PE, particularly the more severe type associated with preterm delivery.

In contrast, the association between greater fetal *HLA-DQB1* heterozygosity and elevated PE risk suggests that fetal *HLA* class II diversity may be associated with adverse immunological effects under specific conditions, potentially by promoting heightened fetal immune activation. Although fetal trophoblasts generally lack expression of classical *HLA* class I and II molecules to limit maternal immune recognition, class II *HLA* expression—including *HLA-DR*^78,79^ and *HLA-DQB1*^80^—has been reported in select fetal immune cell populations within the placenta, particularly resident macrophages (Hofbauer cells) in the villous stroma. Increased class II heterozygosity could expand the repertoire of antigens presented within fetal tissues, thereby amplifying inflammatory signaling and disrupting immune homeostasis at the maternal–fetal interface. Such immune perturbations could contribute to placental inflammation, endothelial dysfunction, and impaired vascular development—key pathogenic mechanisms of PE.

Notably, the strongest fetal heterozygosity effects were observed in preterm PE, which is consistent with the greater contribution of placental and immunologic dysfunction to early-onset disease. Prior studies in Chinese and Dutch cohorts have similarly reported increased fetal heterozygosity at *HLA-A*^41^ and *HLA-DQB1*^23^ in early-onset and severe PE; our findings replicate and extend these observations in a substantially larger, multi-ancestry cohort. It remains possible that the observed *HLA-DQB1* association reflects linkage disequilibrium with nearby regulatory or non-classical *HLA* genes, such as *HLA-G*,^81^ which is expressed on EVT and plays a key role in modulating maternal NK-cell responses. Disentangling locus-specific effects from linked immunoregulatory variation will require deeper genomic characterization of the *HLA* region together with targeted functional studies.

Our maternal–fetal *HLA* mismatch analyses revealed limited overall associations with PE, consistent with the mixed or largely null findings reported in prior studies examining maternal–fetal *HLA* matching and PE risk.^22,26–28^ Most earlier investigations evaluated only a subset of classical *HLA* loci (typically *HLA-A*, *-B*, *-DR*, and *-DQ*), included small numbers of PE cases, and did not examine PE subtypes. Notably, a recent Dutch study that incorporated *HLA-C*—the only classical class I *HLA* gene expressed by fetal trophoblasts—reported lower *HLA-C* mismatch in PE compared with uncomplicated pregnancies.^28^ In this context, our observation that maternal–fetal *HLA-C* mismatch is associated with reduced risk of preterm PE suggests that maternal–fetal compatibility at specific *HLA* loci, rather than overall matching or mismatch burden, may influence PE susceptibility.

Although fetal trophoblasts do not typically express *HLA* class II molecules to prevent maternal immune rejection, aberrant expression of class II proteins, including *HLA-DR* and *HLA-DQ*, in placental syncytiotrophoblasts has been reported under inflammatory conditions, including PE.^82,83,84^ Accordingly, the observed association between *HLA-DQA1* mismatch and increased risk of preterm PE may reflect dysregulated class II–mediated immune activation in the placenta. This aberrant placental class II *HLA* expression may arise as part of an inflammatory response, as class II gene expression is controlled by the Class II Transcription Activator (CIITA), which is induced by pro-inflammatory cytokines such as interferon gamma (IFNG).^85^ Although the downstream pathophysiologic consequences remain undefined, this dysregulation may promote inappropriate CD4⁺ T-cell activation at the maternal–fetal interface. Thus, these findings reinforce the concept that locus-specific and context-dependent maternal–fetal *HLA* interactions, rather than overall mismatch burden, shape susceptibility to PE.

Taken together, these maternal and fetal effects underscore a finely tuned immunologic balance required for a successful pregnancy. Maternal *HLA* diversity may promote immune tolerance and adaptability, whereas greater fetal *HLA* diversity—particularly within class II genes—may be associated with increased susceptibility to immune-mediated pathology. This compartment-specific pattern likely reflects distinct immunologic pressures acting on the maternal and fetal genomes during pregnancy, consistent with emerging co-evolutionary^86^ and cooperative and competitive frameworks of cell interaction and immune signaling at the maternal–fetal interface.^87,88^ These findings highlight the importance of considering maternal and fetal genomes separately when evaluating immunogenetic contributions to PE risk.

The absence of significant associations with individual *HLA* alleles is consistent with large-scale PE GWAS, which have identified few robust signals within the *HLA* region.^24,25^ Together with our heterozygosity and maternal–fetal mismatch analyses, this convergence of evidence suggests that PE risk is unlikely to be driven by single *HLA* alleles, but rather by polygenic and structural features of *HLA* diversity that shape immune balance at the maternal–fetal interface. Our largely null findings for global *HLA* compatibility further argue against simple matching models and underscore the need for more nuanced frameworks that incorporate locus-specific effects, placental expression patterns, and cellular context—including interactions with maternal *KIR* genotypes and the contribution of non-classical *HLA* genes such as *HLA-G* and *HLA-E*.

The inclusion of both US and Colombian participants strengthens the generalizability of our findings. Despite substantial differences in allele frequencies between cohorts, *HLA* heterozygosity patterns were highly conserved—exceeding 80% for all loci except *HLA-DPA1* in both populations—and allele frequencies were strongly correlated, reflecting the evolutionary stability of *HLA* diversity. These observations underscore the value of multi-ancestry study designs for interrogating highly polymorphic genomic regions such as *HLA*.

Our findings also have potential translational relevance. Incorporating maternal and fetal *HLA* diversity into predictive models, alongside clinical factors and angiogenic biomarkers, may improve risk stratification for PE, particularly for early and preterm subtypes. More broadly, these results motivate integrative approaches that combine immune gene profiling with functional immune effector assays to elucidate the molecular mechanisms underlying immune activation and placental dysfunction in PE and to identify candidate pathways for therapeutic intervention.

This analysis has several limitations. Although we used a validated multi-ancestry imputation panel, imputation performance may vary across populations. Sample size limitations, particularly in fetal and maternal–fetal pair analyses, may have reduced power to detect ancestry-specific or interaction effects. In addition, we did not evaluate non-classical *HLA* genes including *HLA-G* and *HLA-E* or *KIR* loci, which are critical components of maternal–fetal immunogenetic interactions. Future studies should integrate non-classical *HLA* and *KIR* genotyping, placental expression data, and functional assays of trophoblast–immune cell interactions to elucidate underlying mechanisms. Combining immunogenetic data with proteomic, transcriptomic, and single-cell analyses will further clarify how *HLA* diversity shapes placental immune environments and affects PE development. Replication in additional multi-ethnic cohorts will be essential to confirm generalizability.

In summary, this study provides evidence for distinct and opposing maternal and fetal associations of *HLA* heterozygosity with PE, supporting a model in which maternal *HLA* diversity enhances immune adaptability and tolerance, while fetal class II diversity may predispose to immune dysregulation, particularly in preterm PE. These findings advance our understanding of maternal–fetal immunogenetics in PE and provide a foundation for future mechanistic and translational studies that integrate genetic insights with functional and molecular profiling to delineate the immunogenetic landscape of PE and identify novel targets for improved early prediction and intervention.

## Data Availability

The data that support the findings of this study are available on request from the corresponding author and/or senior authors, subject to institutional and ethical approvals. Whole-genome sequencing data from TOPMed are available through controlled access via the database of Genotypes and Phenotypes (dbGaP).

## Acknowledgements

Molecular data for the Trans-Omics for Precision Medicine (TOPMed) program was supported by the National Heart, Lung and Blood Institute (NHLBI). Genome Sequencing for “NHLBI TOPMed: The Boston-Colombia Collaborative for Adverse Pregnancy Outcomes” (phs003331.v1.p1, phs003332.v1.p1, phs003333.v1.p1, phs003334.v1.p1, phs003335.v1.p1) was performed at the Broad Institute Genomics Platform (3R01HL092577-06S1, 3U54HG003067-12S2). Core support including centralized genomic read mapping and genotype calling, along with variant quality metrics and filtering were provided by the TOPMed Informatics Research Center (3R01HL-117626-02S1; contract HHSN268201800002I). Core support including phenotype harmonization, data management, sample-identity QC, and general program coordination were provided by the TOPMed Data Coordinating Center (R01HL-120393; U01HL-120393; contract HHSN268201800001I). We gratefully acknowledge the studies and participants who provided biological samples and data for TOPMed.

The analytical work performed in this study was supported by the NIH/NHLBI grant R01 HL163234 (R.S., K.J.G.) and the NIH/Westat TOPMed Fellowship (J.H.). Whole genome sequencing of BCC-PREG samples was supported by the NHLBI TOPMed program under grant X01 HL153753 (K.J.G., J.P.C., R.S.). Creation of the BCC-PREG cohort was supported by a PJP grant from the Preeclampsia Foundation (K.J.G., R.S.), K08 HL146963 (K.J.G.), and R03 HL162756 (K.J.G.).

We are grateful to Dr. Saori Sakaue (University of Washington) for her expert guidance on *HLA* imputation using the multi-ancestry HLA reference panel on the Michigan Imputation Server.

## Web resources

HLA Imputation on Michigan Imputation Server, https://imputationserver.sph.umich.edu/

Common, Intermediate and Well-Documented (CIWD) HLA Alleles in World Populations, https://www.ihiw18.org/component-immunogenetics/download-common-and-well-documented-alleles-3-0/

---

**Figure S1.**
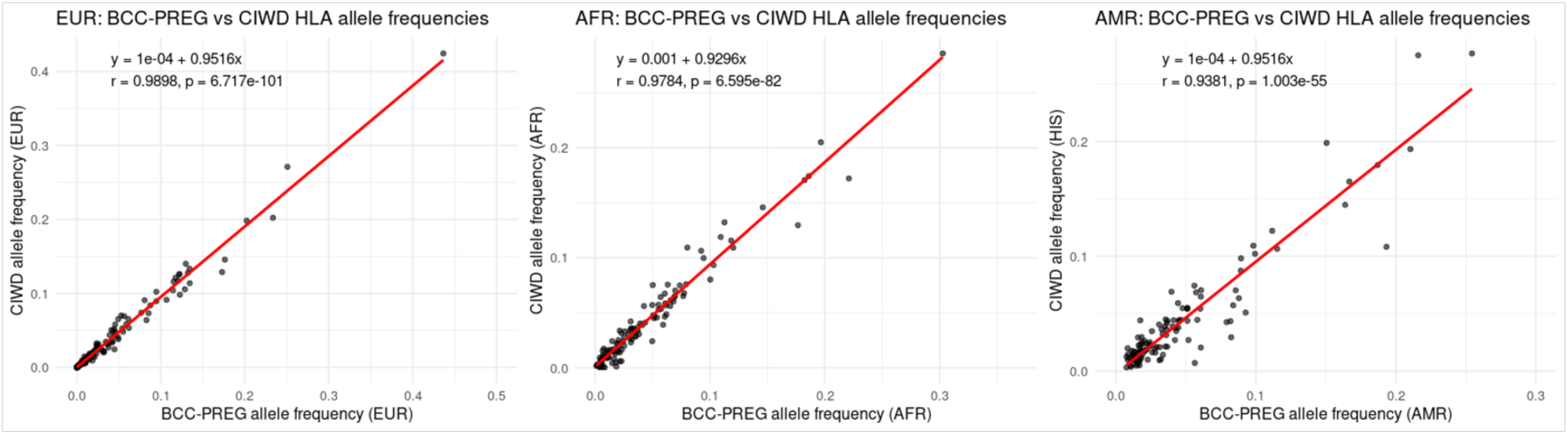
Concordance between *HLA* allele frequencies in BCC-PREG and CIWD reference populations. Scatter plots show the relationship between ancestry-specific *HLA* allele frequencies in the BCC-PREG study and corresponding alleles frequencies in the CIWD *HLA* allele database (n=119 alleles) for European (EUR), African (AFR), and Admixed American (AMR) populations. Because CIWD does not define an AMR category, allele frequencies from the Hispanic (South or Central America/Hispanic/Latino) reference group were used as comparison for BCC-PREG AMR samples. Each point represents one *HLA* allele. Red lines indicate fitted linear regression models, and annotations report Pearson correlation coefficients (r) and corresponding p-values. Correlations were calculated across n = 119 alleles within each ancestry.

